# Seroprevalence and associated factors of maternal cytomegalovirus in Southern Ethiopia

**DOI:** 10.1101/2021.03.03.21252804

**Authors:** Mengistu Hailemariam Zenebe, Zeleke Mekonnen, Eskindir Loha, Elizaveta Padalko

## Abstract

**Introduction:** Cytomegalovirus (CMV) is the most common infection during pregnancy that poses the risk of congenital CMV infections (cCMV) worldwide. The aim of this study was to assess the seroprevalence and associated factors of CMV among pregnant women in Southern Ethiopia.

**Method:** A cross-sectional study was conducted on consecutive women attending the delivery ward at Hawassa comprehensive and specialized hospital. Blood samples collected at the time of delivery were tested for CMV IgG and IgM using ELISA. Study participants responded to a questionnaire about obstetric history and socio-demographic characteristics. Data analysis was performed using SPSS version 20.0 software. Chi-square, bivariable and multivariable logistic regression were used to identify characteristics independently associated with the sero-status of CMV.

**Results:** Seropositivity for CMV IgM antibodies was 8.2% (49/600) (95% CI: 6 −10.5%), whereas the CMV IgG was 88.6% (532/600), (95% CI: 89.5 − 94.0%). Seroprevalence was higher in women of older age, currently unmarried and having nursery schooled children. Moreover CMV seropositivity was significantly associated with any of detected curable STIs. Seroprevalence was not significantly related to previous adverse pregnancy outcome, gravidity, being a child day care occupant mother, and birth weight of the newborn.

**Conclusion:** In the present study, we identified a high rate of CMV IgM seropositivity among pregnant women in southern Ethiopia. Given that there is no existing CMV diagnosis, special attention should be designed for pregnant women in parallel to the existing antenatal care facility. Besides, training health care professionals will support awareness conception among pregnant women concerning the sequels of CMV infection during pregnancy.

**Key questions:** *What is already known?:*

- Cytomegalovirus is the most common infection during pregnancy that can cause congenital CMV infections and known to cause long-term sequelae including sensorineural hearing loss of the developing fetus.
- However, data on maternal CMV infection lacking in Ethiopia besides there is no maternal CMV diagnostics and screening service for pregnant women.

*What are the new findings?:*

- A high prevalence 8.3% of CMV IgM and 88.6% of CMV IgG documented.
- Seroprevalence associated with older age women, currently unmarried and having nursery schooled children in the household. Also CMV seropositivity was significantly associated with any of detected curable STIs.

*What do the new findings imply?:*

- Known that there is no existing maternal CMV screening in Ethiopia, understanding the burden and the effect of maternal CMV infection will offer important information to the health care providers to prevent a sequel to the developing fetus.
- The current study provides valuable information on the associated factors with maternal CMV infection hence, training health care professionals will support awareness conception among pregnant women concerning the sequels for CMV infection during pregnancy.

## Introduction

Cytomegalovirus (CMV) is the most common infection during pregnancy that poses the risk of congenital CMV infections (cCMV) worldwide.(1) In immunocompetent hosts, primary CMV infection may be asymptomatic or may cause mild self-limiting disease with fever, headaches, and myalgia and after primary infection the virus remains latent. Latency following a primary infection may relapse by periodic reactivations that give rise to recurrent infections later in life when the body immunity is suppressed.(2)

CMV infection or reactivation during pregnancy is mostly asymptomatic, however may lead to fetal infection and cCMV syndromes.(3) Congenital CMV infection of the fetus of mothers having pre-existing anti-CMV antibodies is also possible due to risk of reactivation or reinfection with a different strain of CMV during pregnancy.(4) Therefore unlike previous perception the high maternal CMV seroprevalence in developing countries like Ethiopia does not eliminate the threat of cCMV infection of the newborn. Worldwide cCMV following nonprimary maternal infections is more common in individuals of lower socioeconomic background.(5)

So far studies have shown maternal seropositivity rates of previous infection ranging from low (50 to 70%) in developed countries, to high (>□70%) in developing countries.(1) Presently, data on the prevalence of maternal CMV and associated risk factors are scanty in Ethiopia. The only available study conducted in Ethiopia had reported the prevalence of 15.5% for IgM and 88.6% for CMV IgG.(6)

In Africa the highest prevalence of CMV IgG was estimated ranging from 72 – 97.5% (7, 8) and of CMV IgM antibodies ranging from 0-15.5%.(9) However, for several reasons CMV infection among pregnant women in Africa have been overlooked.(10) One of the main reasons for +inattention is the perception that being infected in early childhood endures immunity for subsequent infection, so maternal reactivation or reinfection during pregnancy is unlikely to cause severe congenital infection.(11) However, in pregnant women the immune system is somehow suppressed. So ignoring maternal CMV and the subsequent effect of cCMV infection in Africa is short-sighted; furthermore, the possible confounding effects of HIV infection, malnutrition, tuberculosis, and a general higher disease burden of the continent must be taken into account.(12)

The objective of the study was to assess the seroprevalence of CMV among pregnant women and determine associated factors in Southern Ethiopia. Information about maternal the prevalence of CMV and associated risk factors is almost absent in Ethiopia and this study was the first in the southern region. We hope the study will attract health care professional attention and improve antenatal care in this domain. In the meantime it will generate awareness to the community, mainly pregnant women, regarding the consequence of CMV during pregnancy in Ethiopia.

## Methodology

### Study setting and recruitment

From August to October 2020, a cross-sectional study was conducted among pregnant women who came for delivery in the obstetrics ward at Hawassa University Comprehensive and Specialized Hospital (HU-CSH), Ethiopia. Interrelated with the first phase of this project, where the initial 350 pregnant women had been tested for curable STI (*C. trachomatis, N. gonorrhoeae* and *T. vaginalis*) using GeneXpert (Xpert CT/NG and Xpert TV assays, Cepheid, Sunnyvale, California, USA), our manuscript being submitted and under review (manuscript number PONE-D-20-37668). In this second phase of the study, by including those initially enrolled 350 pregnant women, a total of 600 consecutively enrolled pregnant women were participated. Pregnant women were recruited upon delivery regardless of gestational age. A midwife at the obstetric ward provided general information about the study to all pregnant women before recruitment.

### Socio-demographic, obstetric and behavioral data

Trained midwife at the obstetric ward provided general information about the study to pregnant women who came for delivery. Pregnant women agreeing to join in the study were interviewed using a structured questionnaire translated in Amharic, the language spoken by most people in the study area. The translated questionnaire was pre-tested on random mother at antenatal clinic to ensure the validity and feasibility of the questions as conducted in similar studies. Information related to socio-demographic characteristics (e.g., age, marital status, and educational level), obstetric history, and behavioral data were collected.

### Sample collection and storage

The midwife-nurse aseptically collected a 3 ml blood sample from each subject. The collected samples were transported to the HU-CMHS microbiology laboratory within 12 hours of collection and the processed serums stored at −20 C^0^ until analysis.

### Laboratory methods

Testing was performed in Belgium, Ghent University hospital, department of laboratory medicine, using a commercially available enzyme immunoassay (ELISA) kit (EUROIMMUN Medizinische Labordiagnostika AG, Lübeck, Germany) for anti-CMV IgG and IgM according to the manufacturer’s instructions. Results were evaluated semi-quantitatively by calculating a ratio of the extinction value of the patient sample over the extinction value of the calibrator optical density at 450 nm. Seropositivity was defined according to the guidelines given by the manufacturer, CMV negative when the ratio cut off value was < 0.8, borderline between 0.8 and 1.1, and positive if >1.1 for both IgG and IgM.

### Ethical considerations

Ethical approval from all of the appropriate institutional review boards was obtained. The ethics review committee of Hawassa University (CMHS/283/2012), Jimma University (IHRPGD/458/2020), National Health Research Ethics Review Committee (SRA/14.1/ 144483/2020) Ethiopia, and Ghent University (PA2019-038/BC-08458) Belgium, approved the study. All participants provided written, informed consent for study participation.

### Data analysis

Descriptive statistics were used to characterize the socio-demographic and obstetric and medical characteristics of the participants. We evaluated the seroprevalence of CMV and associated factors using a logistic regression model. Bivariate comparisons using Chi-square or Fisher’s exact test where suitable were used to examine the relationships between participant characteristics and CMV test result. Finally, multivariable logistic regression was used to identify characteristics independently associated with sero-status of CMV and adjusting for other factors. Variables with a significant level of ≤0.2 were included in the final model. P value < 0.05 is considered statistically significant. SPSS software version 20.0 (SPSS Inc. Chicago, IL, USA) was used for all analyses.

## Results

Six hundred pregnant women were assessed for seroprevalence of CMV. The mean of maternal age was 27.0 ± (SD) 5.2, with a ranging between 17 and 41. More than one-third of the study participant were under the age of 25. About one-fourth of the women were Primigravida. Out of the 600 participant 84 (14%) were currently unmarried, 475 (79.2%) were residing in urban setting, 377 (62%) were above or at secondary level of education. Forty-eight (8%) of the newborns were under-weighted (<2.5 Kg), and 64 (10%) of the births were preterm. Regarding STIs test result, 51 (14.6%) pregnant women were tested positive for any of curable STIs (Table 1).

**Table 1.** Maternal characteristics and associated factor with CMV IgM seropositivity in Southern Ethiopia.

| **Characteristics** | **Total (N=600)**<br>**n (%)** | **IgM-Positive**<br>**(n=49) n (%)** | **IgM-Negative**<br>**(n=551) n (%)** | **p-value** |
| --- | --- | --- | --- | --- |
| **Age of mothers (years)** |  |  |  |  |
| <25 | 233 (38.8) | 14 (28.6) | 219 (39.7) | 0.148 |
| 25-29 | 162 (27.0) | 14 (28.6) | 148 (26.9) |  |
| 30- 35 | 170 (28.3) | 15 (30.6) | 155 (28.1) |  |
| >35 | 35 (5.8) | 6(12.2) | 29 (5.3) |  |
| **Marital status** |  |  |  |  |
| Married | 516 (86.0) | 37 (75.5) | 479 (86.9) |  |
| Currently unmarried | 84 (14.0) | 12 (24.5) | 72 (13.1) | 0.027 |
| **Residence** |  |  |  |  |
| Urban | 475 (79.2) | 44 (89.8) | 431 (78.2) | 0.056 |
| Rural | 125 (20.8) | 5 (10.2) | 120 (21.8) |  |
| **ANC follow up during pregnancy** |  |  |  |  |
| Yes | 576 (96.0) | 48 (98.0) | 528 (95.8) |  |
| no | 24 (4.0) | 1 (2.0) | 23 (4.2) | 0.712 |
| **Employed as daycare worker** |  |  |  |  |
| Yes | 40 (6.7) | 6 (12.2) | 34 (6.2) | 0.126 |
| No | 560 (93.3) | 43 (87.8) | 517 (93.8) |  |
| **Employed as health care** |  |  |  |  |
| Yes | 32 (5.3) | 6 (12.2) | 26 (4.7) | 0.055 |
| no | 568 (94.7) | 43 (87.8) | 525 (95.3) |  |
| **Education** |  |  |  |  |
| Primary and below | 223 (37.2) | 13 (26.5) | 210 (38.1) | 0.108 |
| Secondary and above | 377 (62.8) | 36 (73.5) | 341 (61.9) |  |
| **Gestational age** |  |  |  |  |
| Term | 536 (89.3) | 36 (73.5) | 500 (90.7) |  |
| Preterm | 64 (10.7) | 13 (26.5) | 51 (9.3) | <0.001 |
| **Birth weight** |  |  |  |  |
| <2.5 Kg | 48 (8.0) | 3 (6.1) | 45 (8.2) | 0.789 |
| >2.5 Kg | 552 (92.0) | 46 (93.9) | 506 (91.8) |  |
| **Gravidity** |  |  |  |  |
| Primigravida | 147 (24.5) | 14 (28.6) | 133 (24.1) | 0.489 |
| Multigravida | 453(75.5) | 35 (71.4) | 418 (75.9) |  |
| **Previous adverse pregnancy outcome \*** |  |  |  |  |
| Yes | 39 (8.6) | 2 (5.7) | 37 (8.9) | 0.756 |
| No | 414 (91.4) | 33 (94.3) | 381 (91.1) |  |
| **Knowledge on congenitally transmitted infections** |  |  |  |  |
| Yes | 25 (4.2) | 1 (2.0) | 24 (4.4) |  |
| No | 575 (95.8) | 48 (98.0) | 527 (95.6) | 0.712 |
| **Under-five children in the household** |  |  |  |  |
| Yes | 396 (66.0) | 31 (63.3) | 365 (66.2) | 0.673 |
| no | 204 (43.0) | 18 (36.7) | 186 (33.8) |  |
| **Daycare or Nursery school baby in the household** |  |  |  |  |
| Yes | 259 (43.2) | 31 (63.3) | 228 (41.4) | 0.003 |
| no | 341 (56.8) | 18 (36.7) | 323 (58.6) |  |
| **Sharing feeding cup with children** |  |  |  |  |
| Yes | 107 (17.8) | 14 (28.6) | 93 (16.9) | 0.040 |
| no | 493 (82.2) | 35 (71.4) | 458 (83.1) |  |
| **Sharing eating utensil with children** |  |  |  |  |
| Yes | 88 (14.7) | 8 (16.3) | 80 (14.5) | 0.732 |
| No | 512 (85.3) | 41 (83.7) | 471 (85.5) |  |
| **Sharing teeth brush with children** |  |  |  |  |
| Yes | 42 (7.0) | 3 (6.1) | 39 (7.1) | 0.999 |
| No | 558 (93.0) | 46 (93.9) | 512 (92.9) |  |
| ***N.gonorrhoeae* detected (n=350)** |  |  |  |  |
| Yes | 15 (4.3) | 3 (10.0) | 12 (3.8) | 0.128 |
| No | 333 (95.7) | 27 (90.0) | 306 (96.2) |  |
| ***C. trachomatis* detected (n=350)** |  |  |  |  |
| Yes | 29 (8.3) | 5 (16.7) | 24 (7.5) | 0.089 |
| No | 319 (91.7) | 25 (83.3) | 294 (92.5) |  |
| ***T. vaginalis* detected (n=350)** |  |  |  |  |
| Yes | 11 (3.1) | 2 (6.9) | 9 (2.8) | 0.241 |
| No | 335 (96.8) | 27 (93.1) | 308 (97.2) |  |
| **Any of curable STI detected (n=350)** |  |  |  |  |
| Yes | 51 (14.6) | 10 (33.3) | 41 (12.8) | 0.005 |
| No | 299 (85.4) | 20 (66.7) | 279 (87.2) |  |

\*previous adverse pregnancy includes; early neonatal death, stillbirth and preterm birth : STI; Sexually transmitted infections

In this study, around 96% of mothers had no knowledge of congenitally transmitted infection or the associated risks in pregnancy and about 9% of them had previous adverse pregnancy outcome. The chi-squire analysis shown that, seropositivity for CMV IgM significantly associated (p<0.05) with marital status, gestational age, having nursery school baby in the household, sharing a cup with children and having any of detected curable STIs. However, there was no significant association with birth weight, gravidity and having previous adverse pregnancy outcome i.e. preterm birth, stillbirths and early neonatal death (Table 1).

### Seroprevalence

Seropositivity of CMV IgM antibodies was 8.2% (49/600) (95% CI: 6 −10.5%), whereas seroprevalence of CMV IgG was 88.6% (532/600), (95% CI: 89.5 − 94.0%). Of 532 CMV IgG positive women, 483 (80.4%) were negative for IgM. Among all pregnant women, 68 (11.4%) were tested negative for both anti-CMV IgG and IgM, and none showed anti-CMV IgG negativity but IgM positivity (Table 2).

**Table 2.** Cytomegalovirus IgM and IgG test result of pregnant women.

| Anti CMV IgM antibody | Anti CMV IgG antibody n (%) |  | Total n (%) |
| --- | --- | --- | --- |
|  | Positive | Negative |  |
| Positive | 49 (8.2) | 0 (0) | 49 (8.2) |
| Negative | 483 (80.4) | 68(11.4) | 551 (91.8) |
| Total | 532 (88.6) | 68(11.4) | 600 |

### CMV seropositivity and associated factors

In bivariable analysis, seropositivity was more common in elder women (>35) compared to the youngest age group, in women who were currently unmarried, giving preterm birth, sharing a feeding cup with children or having nursery schooled children. Moreover women were positive for any of curable STIs also had a higher seroprevalence of CMV compared to those negative for STIs (Table 3).

**Table 3.** Unadjusted and adjusted associated factors of maternal CMV IgM seropositivity in Southern Ethiopia.

| Characteristics | Un adjusted |  | Adjusted |  |
| --- | --- | --- | --- | --- |
|  | OR 95% CI) | P-value | OR (95% CI) | P-value |
| **Age of mothers (years)** |  |  |  |  |
| <25 | 1 |  | 1 |  |
| 25-29 | 1.5 (0.7 -3.2) | 0.318 | 1.2 (0.4 – 4.0) | 0.739 |
| 30- 35 | 1.5 (0.7 -3.2) | 0.283 | 3.0 (1.0 – 9.0) | 0.048 |
| >35 | 3.2 (1.2– 9.1) | 0.026 | 4.9 (1.0 – 23.4) | 0.047 |
| **Marital status** |  |  |  |  |
| Married | 1 |  | 1 |  |
| Currently unmarried | 2.2 (1.1 -4.3) | 0.030 | 3.8 (1.3 -11.2) | 0.015 |
| **Residence** |  |  |  |  |
| Urban | 2.5 (1.0 -6.3) | 0.064 | 2.3 (0.7 -7.9) | 0.171 |
| Rural | 1 |  | 1 |  |
| **Daycare worker** |  |  |  |  |
| Yes | 2.1 (0.8 -5.3) | 0.110 | 1.1 (0.2- 5.4) | 0.857 |
| No | 1 |  | 1 |  |
| **Health care worker** |  |  |  |  |
| Yes | 2.8 (1.1 - 7.2) | 0.031 | 1.2 (0.2 – 7.4) | 0.841 |
| no | 1 |  | 1 |  |
| **Education** |  |  |  |  |
| Primary and below | 0.6 (0.3 -1.1) | 0.111 | 0.7 (0.3 -1.8) | 0.475 |
| Secondary and above | 1 |  | 1 |  |
| **Gestational age** |  |  |  |  |
| Term | 1 |  | 1 |  |
| Preterm | 3.5 (1.8 – 7.1) | <0.001 | 3.9 (1.5 -10.3) | <0.006 |
| **Daycare or Nursery school baby** |  |  |  |  |
| Yes | 2.4 (1.3 – 4.5) | 0.004 | 2.7 (1.1 – 6.4) | 0.027 |
| no | 1 |  | 1 |  |
| **Sharing a cup with children** |  |  |  |  |
| Yes | 2.0 (1.1 – 3.8) | 0.044 | 2.2 (0.9 – 5.4) | 0.074 |
| no | 1 |  |  |  |
| **Any of curable STIs (n=350)** |  |  |  |  |
| Yes | 3.4 (1.5- 7.8) | 0.004 | 4.1 (1.6- 10.6) | 0.003 |
| No | 1 |  |  |  |

Furthermore, in multivariable logistic regression maternal IgM seropositivity was associated with elder age groups (AOR = 4.9, 95% CI: 1.0–23.4), currently unmarried women (AOR = 3.8, 95% CI: 1.7–7.9), preterm birth (AOR = 3.9, 95% CI: 1.5–10.3) and having nursery schooled children (AOR = 2.7, 95% CI: 1.1–6.4). Mothers with STIs has an association with seroprevalence (AOR = 4.1, 95% CI: 1.6–10.6) compared to mothers who were diagnosed negative for STIs.

Maternal seroprevalence was not significantly different in relation to residence, education level, to mother’s occupation; being employed in child daycare or being health care worker (Table 3).

## Discussion

In this study an overall 8.2% CMV IgM and 88.7% seroprevalence of CMV IgG were detected among pregnant women in southern Ethiopia. The associated factors of seropositivity were age, marital status, the presence of a curable STIs and sharing a cup with children. A statistically significant association was also observed between CMV seropositivity and preterm delivery.

The reported seropositivity of 88.6% CMV IgG in this study was comparable to a previous report of 88.5% h in central Ethiopia but a substantially higher rate (15.5%) of CMV IgM was detected in this study.(6) Seropositivity of CMV IgG in our study was in line with a review done in Africa, with ranges from 60% to 100%.(9) Seropositivity rates of 77.3% for IgG and 8.1% for IgM in Kenya,(7) 93% for IgG and 11.1% for IgM in Nigeria,(13) 94% for IgG and 8.5% for IgM in Tanzania(14) were comparable to our finding.

In our study seroprevalence of CMV IgM is in concordance with several African studies.(7, 13-15) However, our rate was higher when compared to 0.4% in Tanzania,(16) 2.5% in Sudan,(17), and 7% in Egypt.(18) In the absence of maternal screening this high rate is alarming for policy makers. By far developed countries pregnant women are screened for CMV because of the consequence for the fetus and newborn.(19, 20) However, in most developing countries including Ethiopia, maternal CMV still lack awareness, overlooked and not diagnosed at least for pregnant women.(21) The high rate of positive CMV IgM may not only reflect primary infection but might also be attributed to reinfection or reactivation of CMV during pregnancy. So the reported high seropositivity in this study point to the existing negligence and the need to start screening to detect pregnant women at risk for congenital transmission of CMV.

In this study, elder age has significant association with seroprevalence. The same finding was reported in Sudan,(17) Kenya,(22), and Tanzania.(16) In addition a large scale study in London also reported that seropositivity increases with increasing age.(23) But, in a study in Egypt(18) and in Nigeria(24) age had no association with maternal CMV infection. The highest seroprevalence with age may indicate the more exposure of elders than youngsters or might be the existence of previous infection which can probably reactivated in the current pregnancy.

On the other hand, significant association of seroprevalence with having nursery schooled children among household was observed. For pregnant women the predictable source of CMV infection is young children mainly exposure to nursery schooled children.(25) Children easily get infected in school and frequently shed CMV in their saliva or urine for many years continuously where spreads readily in preschool setting.(26) This places seronegative pregnant women who have a young child in the home or in day care at increased risk of seroconversion.(27, 28) Susceptibility to acquisition of CMV infection is high possibly through the direct contact with contagious secretions from their own children essentially in situation of poor hygienic practice like in Ethiopia.(23)

Among candidate predictors for maternal CMV seropositivity, occupation like being health care worker, or child day care worker; being multigravida, lower educational level and having other children at home were not detected. However, significant association with women who delivered preterm was reported. Although, preterm delivery might be due to the effect of maternal CMV infection we can not to say the risk factor as there are a lot of other confounding factors for preterm.(29)

Likewise, seropositivity was found to be significantly associated with STIs detected at delivery and currently being unmarried. Mothers who were positive for any of curable STIs had a four-time CMV seropositivity. It is also reported that STIs including CMV is more common in unmarried pregnant women.(23, 25, 30) Although cytomegalovirus is a virus that is transmitted through many body fluids, sexual transmission from a seropositive male partner is an additional established route by which women may be infected with CMV.(31) Indeed, it is somehow expected that sexual transmission is also responsible for reinfection of seropositive mothers with different virus strains in high-seroprevalence populations.(32)

The limitations of the present study are that we lack differentiation of CMV IgM positives of either primary or secondary (reinfection or reactivation) since we collected samples at the end of pregnancy and avidity test will not suitable. Moreover, it was a hospital-based study and not representative of all pregnant mothers in the locality since a significant portion of mothers may not deliver in the hospital. We lack also appropriate risk factors assessment tool as we mainly focused on seroprevalence and nature of the enrolled participant (time of labour), hence more representative large scale survey is needed to identify possible risk factors prospectively.

## Conclusion

In the present study, we documented a high rate of CMV seroprevalence among pregnant women in southern Ethiopia. The presence of a curable STIs, elder age, unmarried women, and having nursery schooled children have a significant association with seropositivity. Given that there is no existing CMV diagnostic facility, special attention should be designed for pregnant women in parallel to the existing antenatal care service. Besides, training health care professionals will support awareness conception for pregnant women concerning the sequels of CMV infection during pregnancy.

## Data Availability

All data available in the main document

## Acknowledgments

We thank the HU-CSH microbiology laboratory staffs for provision of all laboratory accommodations during sample processing and storage. We would like to recognise the study participants and a special thanks to midwife nurses at the obstetrics ward of the HU-CSH. Lastly, we want to express our thanks to VLIR-UOS for financial support.

## Author Contributions

MH and EP contributed to the study design and conceptualisation. MH carried out the laboratory work. MH, EL, ZM and EP performed the statistical analysis and interpretation. All authors provided critical review and contributed to the write-up and approved the final version of the manuscript. MH had final responsibility to submit for publication. All authors read and amended drafts of the paper and approved the final version.

## Patient consent

Obtained. All participants provided written, informed consent for study participation.

## Ethics approval

Ethical approval from all of the appropriate institutional review boards was obtained. The ethics review committee of Hawassa University (CMHS/283/2012), Jimma University (IHRPGD/458/2020), National Health Research Ethics Review Committee (SRA/14.1/ 144483/2020) Ethiopia, and Ghent University (PA2019-038/BC-08458) Belgium, approved the study.

## Data sharing statement

No additional data are available.

